# Air pollution, SARS-CoV-2 transmission, and COVID-19 outcomes: A state-of-the-science review of a rapidly evolving research area

**DOI:** 10.1101/2020.08.16.20175901

**Authors:** A Bhaskar, J Chandra, D Braun, J Cellini, F Dominici

## Abstract

**Background:** As the coronavirus pandemic rages on, 692,000 (August 7, 2020) human lives and counting have been lost worldwide to COVID-19. Understanding the relationship between short- and long-term exposure to air pollution and adverse COVID-19 health outcomes is crucial for developing solutions to this global crisis.

**Objectives:** To conduct a scoping review of epidemiologic research on the link between short- and long-term exposure to air pollution and COVID-19 health outcomes.

**Method:** We searched PubMed, Web of Science, Embase, Cochrane, MedRxiv, and BioRxiv for preliminary epidemiological studies of the association between air pollution and COVID-19 health outcomes. 28 papers were finally selected after applying our inclusion/exclusion criteria; we categorized these studies as long-term studies, short-term time-series studies, or short-term cross-sectional studies. One study included both short-term time-series and a cross-sectional study design.

**Results:** 27 studies of the 28 reported evidence of statistically significant positive associations between air pollutant exposure and adverse COVID-19 health outcomes; 11 of 12 long-term studies and all 16 short-term studies reported statistically significant positive associations. The 28 identified studies included various confounders, spatial and temporal resolutions of pollution concentrations, and COVID-19 health outcomes.

**Discussion:** We discuss methodological challenges and highlight additional research areas based on our findings. Challenges include data quality issues, ecological study design limitations, improved adjustment for confounders, exposure errors related to spatial resolution, geographic variability in testing, mitigation measures and pandemic stage, clustering of health outcomes, and a lack of publicly available data and code.

## Introduction

The COVID-19 pandemic is devastating human health in a manner unseen since the last century. As the pandemic rages on, as of August 7, 2020, the world faces a total of 7.94M confirmed cases and the loss of 692,000 human lives and counting (Wikipedia 2020). The death toll in the United States continues to rise and has even surpassed the US death toll during World War II (Johns Hopkins University and Medicine).

An enormous amount of scientific literature has provided strong evidence that short- and long-term exposure to air pollution leads to an increased risk of mortality and morbidity (US Environmental Protection Agency 2020). Air pollution contributes to respiratory tract infections, chronic obstructive pulmonary disease, coronary heart disease, and diabetes burden, in addition to other chronic conditions (Jiang et al. 2016; Lelieveld and Munzel 2019; US Environmental Protection Agency 2020). A review of evidence supports a relationship between high concentrations of air pollution and the probability of respiratory viruses negatively impacting the respiratory system and exacerbating disease severity (Domingo and Rovira 2020). These health risks are compounded by recent evidence that SARS-CoV-2 can exist on particulate matter, suggesting that air pollution can contribute to viral spread (Setti et al. 2020).

Exposure to air pollutants has been shown to adversely impact respiratory health by inducing oxidative stress, damaging macrophage cells, decreasing the expression of surfactant proteins, and increasing the permeability of lung epithelial cells, all of which can reduce resistance to viral infection (Ciencewicki and Jaspers 2007). In studying outbreaks of SARS, the first human coronavirus, Cui et al. (2003) provided preliminary evidence of a positive association between air pollution and case fatality, although their ecological study did not adjust for confounders.

Severe COVID-19 infection is characterized by a high inflammatory burden, and it can cause viral pneumonia with additional extrapulmonary manifestations and complications including acute respiratory distress syndrome (ARDS) (Chen et al. 2020; Huang et al. 2020; Mehta et al.2020; Ruan et al. 2020; Wang D et al. 2020; Xu Z et al. 2020), which has a mortality rate ranging from 27% to 45% (Diamond et al. 2020). A recent study by our group documented a statistically significant association between long-term exposures to fine particulate matter(PM_2.5_) and ozone and the risk of ARDS among older adults in the United States (Rhee et al. 2019).

To support the scientific community in their work to mitigate risk and develop solutions to address the global COVID-19 crisis, it is crucial to evaluate the evidence to date on the potential associations between exposure to air pollution and COVID-19 health outcomes. Previous reviews of this literature (Benmarhnia 2020; Copat et al. 2020) were limited in scope. This review is unique in its inclusion of both short- and long-term exposure studies as well as a specific focus on identifying unresolved methodological challenges to understanding the association between exposure to air pollution and COVID-19 health outcomes. By chronicling the use of relevant confounders, air pollution datasets, and statistical approaches, we also highlight future opportunities for research on the intersection between air pollution exposure and COVID-19 health outcomes.

## Methods

### Protocols

We conducted this state-of-the science review based on the 5-step process (identification of the research question, identification of relevant studies, selection of studies to include based on inclusion/exclusion criteria, charting of the data, summarization results) outlined by Arksey and O’Malley (Arksey and O’Malley 2005).

### Search strategy

We conducted a literature search of the National Library of Medicine’s PubMed database (National Institutes of Health et al.), Elsevier’s Embase Database, Clarivate Analytics Web of Science Database, Cochrane’s COVID-19 Study Register, Cold Spring Harbor Laboratory’s medRxiv, and Cold Spring Harbor Laboratory’s bioRxiv. The final search date was June 9, 2020. “Air pollution” and “COVID-19” were used as the keywords for the searches, along with their synonyms, including “PM2.5,” “PM10,” “particulate,” “NO2,” “ozone,” “O3,” etc., for air pollution, and “coronavirus,” “SARS-CoV-2,” “SARSCov19,” etc., for COVID-19. The full search queries used for each database are provided in Supplemental Materials Table 1.

### Study eligibility criteria and study selection

Inclusion/exclusion criteria were designed to identify original research describing the effects of air pollution on COVID-19 health outcomes (Figure 1). We included both published and preprint studies due to the quickly evolving nature of the topic. Only studies published in English up until June 9, 2020 were included.

**Figure 1.**
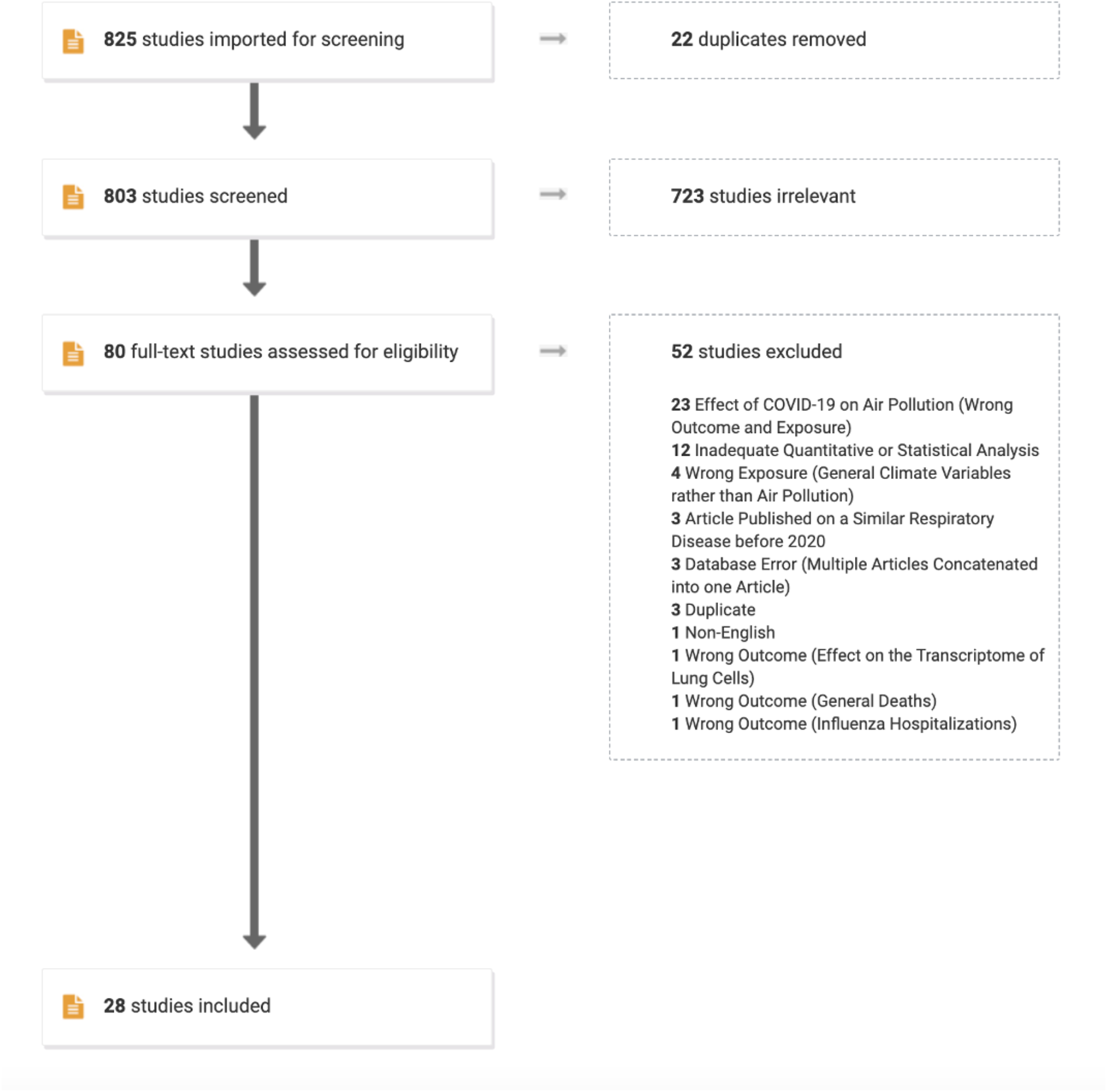
PRISMA figure displaying the screening process for the 825 studies identified in the database searches run on June 9, 2020. In the first round, duplicate studies were removed. In the second round, studies deemed irrelevant by title and abstract review were removed. Finally, studies deemed irrelevant by full-text review were removed, resulting in a total of 28 studies included in the review.

Two screeners (AB and JC) reviewed each identified paper from the database searches in the Covidence online platform. Duplicate papers were removed. The studies were then put through a coarse-grain selection process in which studies were excluded based on a review of the title and abstract. Papers that did not discuss air pollution and COVID-19 health outcomes were excluded. In the second round, full texts were reviewed. Studies that discussed the effects of short- or long-term exposure to air pollution on COVID-19 health outcomes were included. Studies could report various COVID-19 health outcomes, including case incidence, case-fatality rate, mortality rate, case hospitalizations, death rate, and secondary cases produced by an initially infected individual. Studies were neither restricted based on the temporal and spatial resolution of air pollution data or the COVID-19 health data nor on the basis of confounder selections.

Any studies investigating the effects of COVID-19 and the lockdowns caused by COVID-19 on air pollution were excluded as these studies are not the focus of this specific review. Many early studies described the relationship between COVID-19 cases and air pollution without significant quantitative or statistical analysis. These studies concluded that air pollution is related to coronavirus spread because early cases in the pandemic were concentrated in the highly polluted Po Valley and other highly polluted areas of China (Frontera et al. 2020; Martelletti and Martelletti 2020). These studies, which provided only cursory analyses, were also excluded.

### Data extraction

For each selected study, we extracted information about the title, author, and which journal/preprint repository it was housed in, as well as geographical area of study, research question, health outcome, exposure, statistical methods, results, covariates or possible confounders, and code availability.

### Study quality

One study did not describe the statistical methods used but still reported a significant p-value (Baron et al. 2020) and another study used an unspecified air quality metric (Bashir et al. 2020).

## Results

### Selected studies

The search yielded 825 articles across all of the databases (Figure 1). After excluding studies that did not discuss both air pollution and COVID-19, 80 articles were eligible for full text review. Of these, only 28 were full studies published in English that investigated the effects of air pollution on COVID-19 health outcomes. Only 3 of the 28 articles included open access of their code.

Tables 1–3 present a summary of the 28 selected articles. Table 1 summarizes cross-sectional studies that focus on the long-term effects of air pollution on COVID-19 health outcomes (n=12 studies) (Andree 2020; Coccia 2020; Fattorini and Regoli 2020; Liang et al. 2020; Lippi et al. 2020; Ogen 2020; Pansini and Fornacca 2020a, b; Tian H et al. 2020; Tian T et al. 2020; Travaglio et al. 2020; Wu et al. 2020). These studies examined the impact of air pollution exposure over a long period of time prior to the SARS-CoV-2 outbreak. The underlying hypotheses of these studies is that exposure to air pollution over long periods of time negatively impacts respiratory health, thereby increasing the susceptibility to SARS-CoV-2 infection and more severe COVID-19 health outcomes. All of these studies relied on a cross-sectional study design.

**Table 1.**
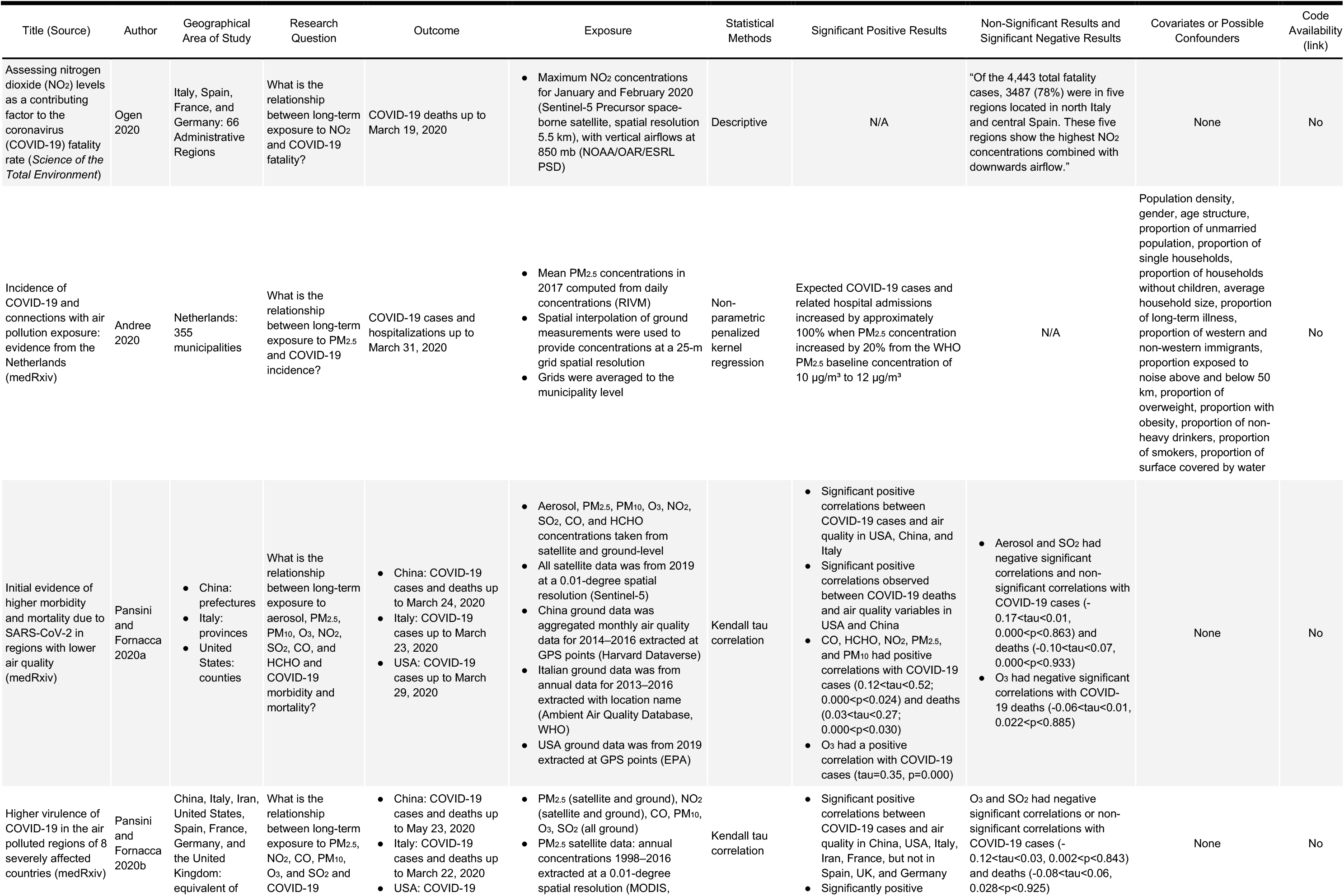
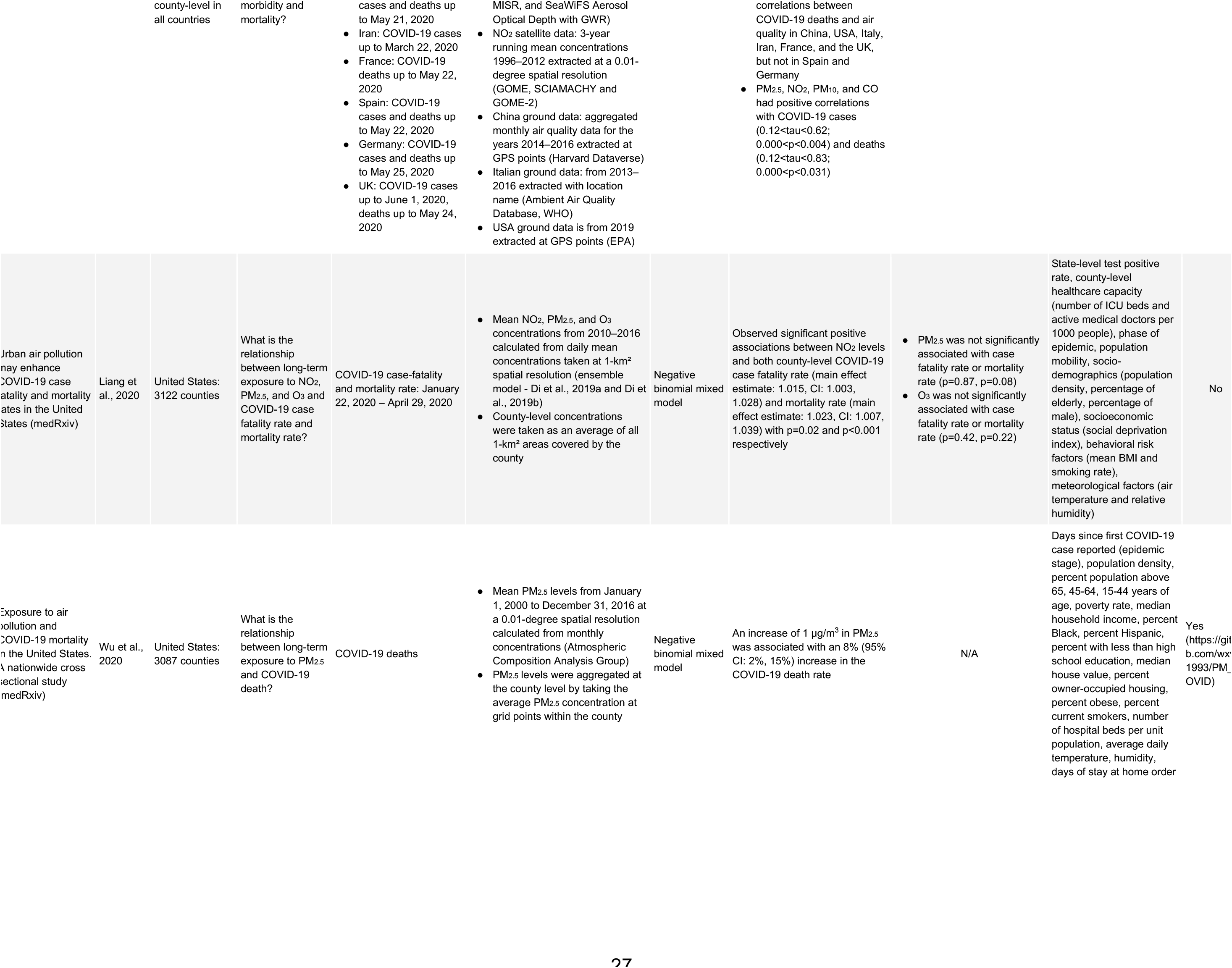
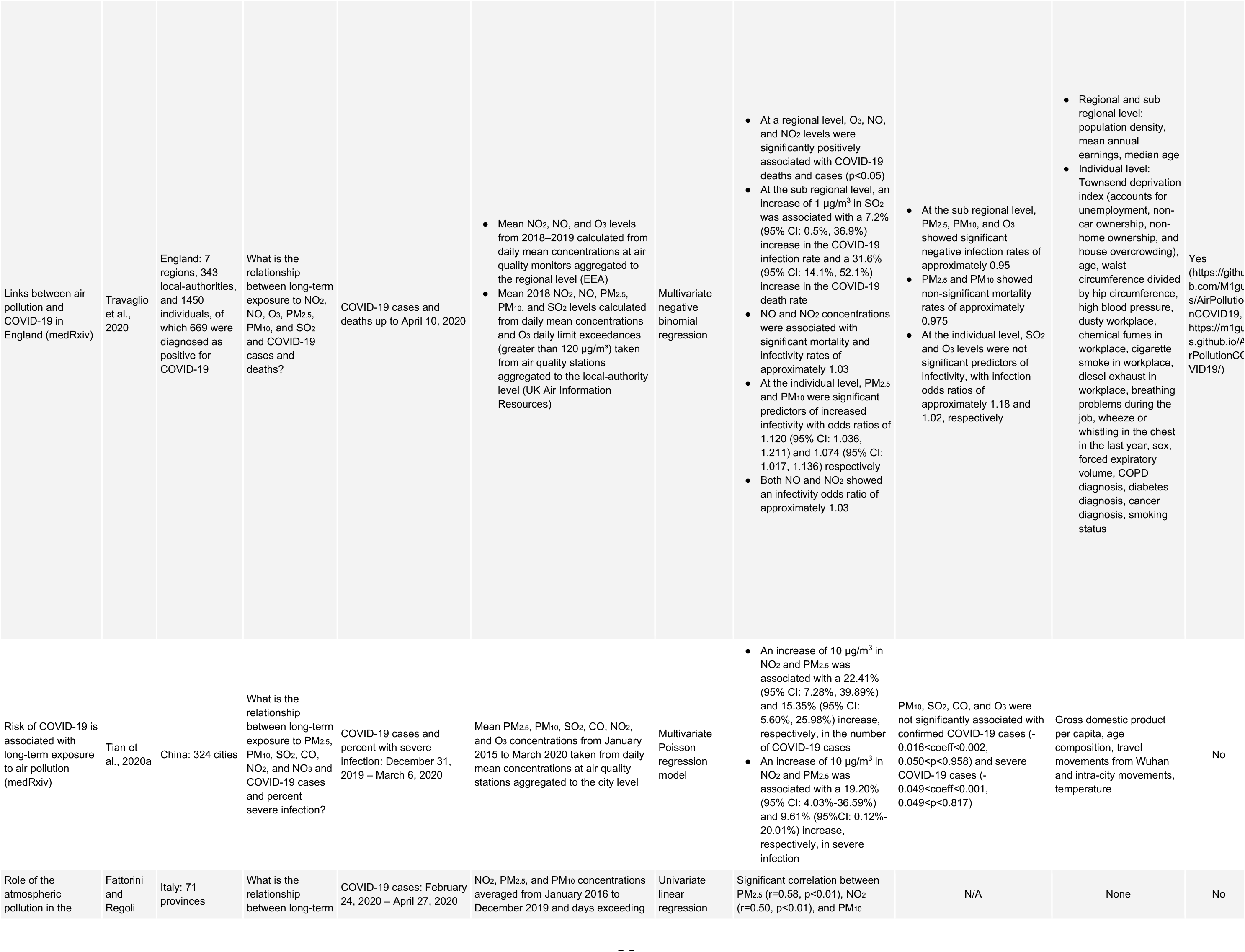
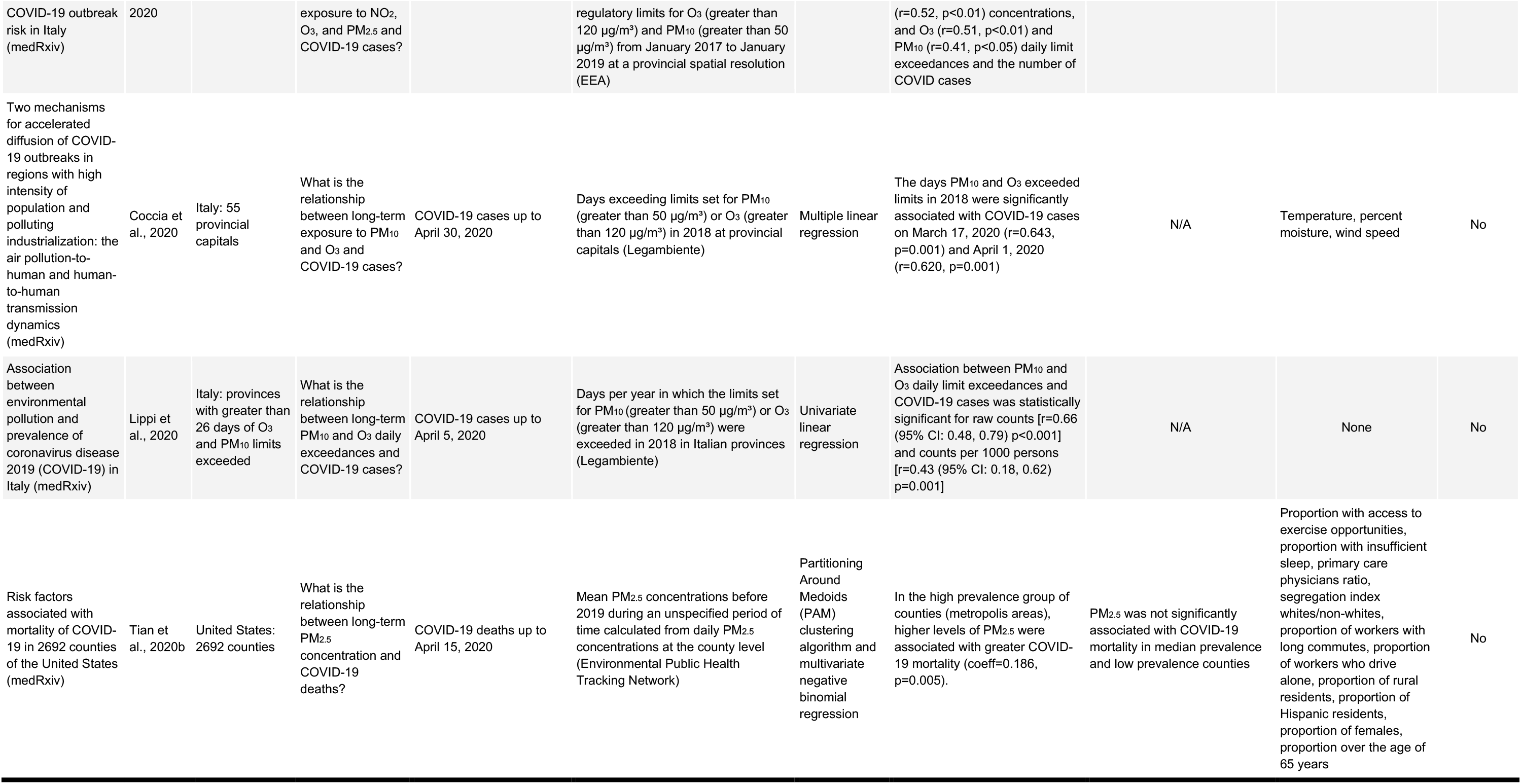
Long-term Studies Included in this Scoping Review

**Table 2.**
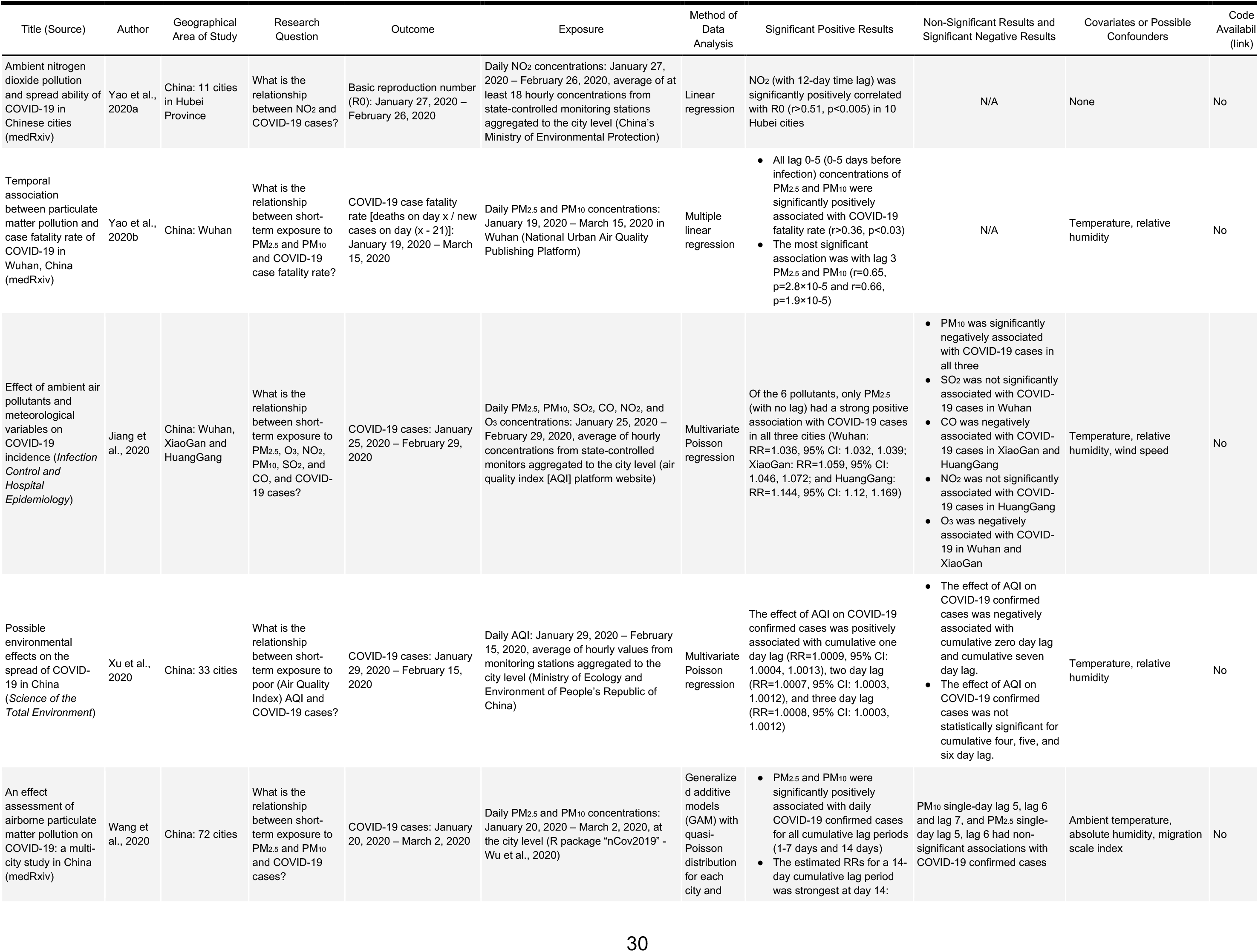
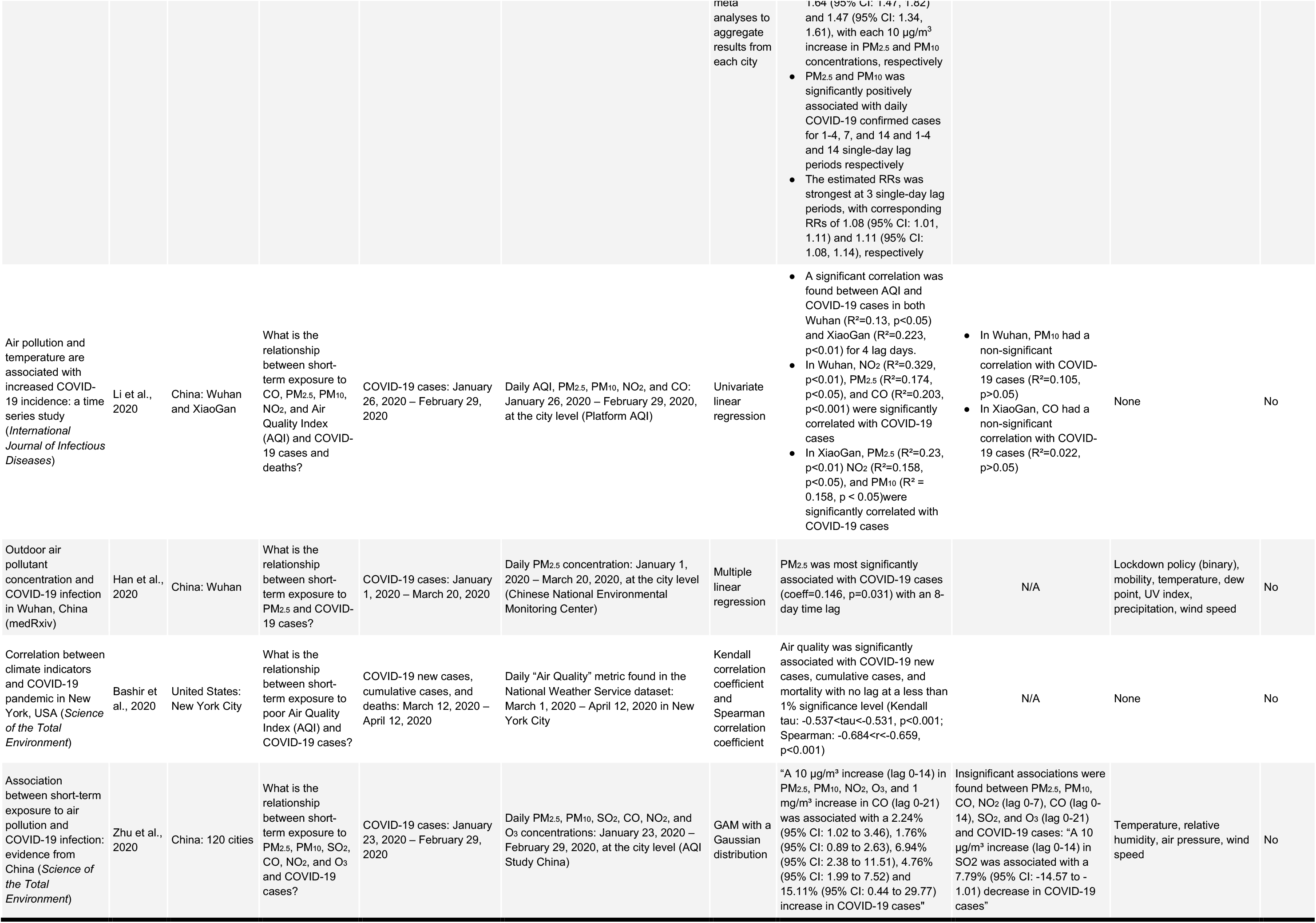
Short-term Time-series Studies Included in this Scoping Review

**Table 3.** Short-term Cross-sectional Studies Included in this Scoping Review

| Title (Source) | Author | Geographical Area of Study | Research Question | Outcome | Exposure | Method of Data Analysis | Significant Positive Results | Non-Significant Results and Significant Negative Results | Covariates or Possible Confounders | Code Availability (link) |
| --- | --- | --- | --- | --- | --- | --- | --- | --- | --- | --- |
| Ambient air pollutants, meteorological factors and their interactions affect confirmed cases of COVID-19 in 120 Chinese cities (medRxiv) | Zhou et al., 2020 | China: 120 cities | What is the relationship between short-term exposure to PM<sub>2.5</sub>, NO<sub>2</sub>, SO<sub>2</sub>, CO, and O<sub>3</sub> and COVID-19 cases? | COVID-19 cases: January 15, 2020 – March 18, 2020 | Mean PM<sub>2.5</sub>, NO<sub>2</sub>, SO<sub>2</sub>, CO, and O<sub>3</sub> concentrations: January 15, 2020 – March 15, 2020, calculated from daily concentrations at the city level (Harvard Dataverse) | Negative binomial regression | CO and PM<sub>2.5</sub> were significantly positively associated with COVID-19 confirmed cases for 0, 3, 7, and 14 lag days (0.000<p<0.045)<br>O<sub>3</sub> was significantly positively associated with COVID-19 confirmed cases for 0 lag days (p=0.045) | NO<sub>2</sub> was not significantly associated with COVID-19 confirmed cases for 0, 3, 7, 14 lag days (0.320 < p < 0.402)<br>O<sub>3</sub> was not significantly associated with COVID-19 confirmed cases for 3, 7, and 14 lag days (0.06 < p < 0.180)<br>SO<sub>2</sub> was significantly negatively associated with COVID-19 confirmed cases for 0, 3, 7, and 14 lag days (p < 0.001) | Average temperature range, diurnal temperature range, relative humidity, wind velocity, air pressure, precipitation, hours of sunshine | No |
| Associations between ambient air pollutants exposure and case fatality rate of COVID-19: a multi-city ecological study in China (medRxiv) | Zhang et al., 2020a | China: 24 provinces and 13 main cities in Hubei Province | What is the relationship between short-term exposure to PM<sub>2.5</sub>, PM<sub>10</sub>, and NO<sub>2</sub> and COVID-19 case fatality rate? | COVID-19 case fatality rate: January 22, 2020 – March 5, 2020 | Cumulative PM<sub>2.5</sub>, PM<sub>10</sub>, and NO<sub>2</sub> concentrations: December 25, 2019 – March 5, 2020, extracted from daily concentrations at air quality monitors aggregated to the provincial and city level (National Air Quality Real-time Publishing System of China) | Multiple linear regression | At the province level, PM<sub>2.5</sub>, PM<sub>10</sub>, and NO<sub>2</sub> were significantly correlated with COVID-19 case fatality rate for 14-day and 28-day lag periods (0.62<r<0.73; 0.000<p<0.002)<br>At the city level in Hubei, NO<sub>2</sub> was significantly correlated with COVID-19 case fatality rate (r=0.697, p=0.008) | At the city level in Hubei, PM<sub>2.5</sub> and PM<sub>10</sub> were not significantly associated with COVID-19 case fatality rate (0.032 < r < 0.185; 0.513 < p < 0.917) | None | No |
| Ambient nitrogen dioxide pollution and spread ability of COVID-19 in Chinese cities (medRxiv) | Yao et al., 2020a | China: 63 cities | What is the relationship between short-term exposure to NO<sub>2</sub> and the spreadability of COVID-19? | Basic reproduction number (R0) using case data up to February 26, 2020 | Mean daily NO<sub>2</sub> concentrations: January 27, 2020 – February 26, 2020, calculated as an average of at least 18 hourly concentrations from state-controlled monitoring stations aggregated to the city level (National Urban Air Quality Publishing Platform) | Multiple linear regression | NO<sub>2</sub> was positively associated with R0 in all cities ($\chi^2=10.18$, p=0.037) | N/A | Temperature and humidity | No |
| Severe air pollution links to higher mortality in COVID-19 patients: the "double-hit" hypothesis (*Journal of Infectious Diseases*) | Frontera et al., 2020 | Italy: 21 regions | What is the relationship between short-term exposure to PM<sub>2.5</sub> and COVID-19 health outcomes? | COVID-19 cases, hospitalizations, ICU admissions, and deaths up to March 31, 2020 | Mean PM<sub>2.5</sub> concentrations in February 2020, calculated from daily levels from measurement stations aggregated to the regional level (Air-Matters App and Website) | Univariate linear regression | PM<sub>2.5</sub> was significantly associated with COVID-19 cases (r=0.64, p=0.0074), ICU admissions per day (r=0.65, p=0.0051), deaths (r=0.53, p=0.032), and hospitalized cases (r=0.62, p=0.0089) | N/A | None | No |
| Spatial correlation of particulate matter pollution and death rate of COVID-19 (medRxiv) | Yao et al., 2020c | China: 49 cities | What is the relationship between short-term exposure to PM<sub>2.5</sub> and PM<sub>10</sub> and COVID-19 deaths? | COVID-19 death rate (deaths/confirmed cases) up to March 22, 2020 | Mean daily PM<sub>2.5</sub> and PM<sub>10</sub> concentrations in March 2020 at the city level (National Urban Air Quality Publishing Platform) | Multiple linear regression | PM<sub>2.5</sub> was positively associated with COVID-19 death rate ($\chi^2=13.10$, p=0.011) and PM<sub>10</sub> ($\chi^2=12.38$, p=0.015) | N/A | Temperature, relative humidity, gross domestic product per capita, hospital beds per capita | No |
| Spatial-temporal variations in atmospheric | Fronza et al., 2020 | France, Germany, Spain: 47 | What is the relationship between short- | Total COVID-19 cases and a binary variable for epidemic escalation: | Mean daily and mean daily maximum concentrations for PM<sub>2.5</sub>, PM<sub>10</sub>, NH<sub>3</sub>, and O<sub>3</sub>: February 10, 2020 – April 10, | Spearman correlation and an | PM<sub>2.5</sub>, PM<sub>10</sub>, and NH<sub>3</sub> showed a significant positive correlation with COVID-19 | O<sub>3</sub> had a significantly negatively correlated with COVID-19 cases per million | Temperature, dew point temperature, wind speed, surface pressure | Yes (https://github.com/CO) |
| factors contribute to SARS-CoV-2 outbreak (*Viruses*) |  | regional capitals<br>Italy: 107 major cities | term exposure to PM<sub>2.5</sub>, PM<sub>10</sub>, NH<sub>3</sub>, and O<sub>3</sub> and COVID-19 cases? | March 25, 2020 – April 10, 2020 | 2020, calculated by aggregating hourly concentrations at the city level | Artificial Neural Network (ANN) to predict epidemic escalation | cases per million persons (0.58<r<0.68)<br>The ANN classifier predicting epidemic escalation with both PM<sub>2.5</sub> and O<sub>3</sub> was significant with respect to the null predictor | persons (-0.5 < r < -0.44) |  | VID19Upcome/Europe) |
| COVID-19 pandemic in relation to levels of pollution with PM<sub>2.5</sub> and ambient salinity. an environmental Wake-up Call (medRxiv) | Baron et al., 2020 | 7 cities with high numbers of COVID-19 cases, including Wuhan, Qom,<br>Bergamo, Paris, Madrid, London and New York<br>8 coastal or small island cities with low numbers of COVID-19 cases, including Seoul, Napoli, Brest, Eastbourne, Taipei, Hong Kong<br>Small countries such as Malta and Cyprus | What is the relationship between short-term exposure to PM<sub>2.5</sub> and COVID-19 cases? | Difference in PM<sub>2.5</sub> concentrations for two groups of cities, delineated by low or high number of COVID-19 cases in an unspecified time period | Minimum and maximum PM<sub>2.5</sub> levels 1 month before and 1 month after statutory lockdown or mandatory restrictions at the city and country level (China National Environmental Monitoring Centre) | Unspecified | "One month prior to the statutory national lock-down or mandatory restrictions, there appear to be high levels of particulate matter, PM<sub>2.5</sub> (min-max 67.4 - 118.7 AQI), in cities/countries which had a high incidence of Covid-19 infection compared to lower levels in cities/countries that have contained the infection (min-max 45.6 - 79.8) (p<0.046)" | N/A | None | No |
| The potential role of particulate matter in the spreading of COVID-19 in northern Italy: first evidence-based research hypotheses (medRxiv) | Setti et al., 2020 | Italy: 110 provinces | What is the relationship between short-term exposure to PM<sub>10</sub> and COVID-19 cases? | COVID-19 cases: February 24, 2020 – March 13, 2020 | Daily PM<sub>10</sub> concentrations exceeding legal limit of 50 µg/m<sup>3</sup>: February 9, 2020 – February 29, 2020, extracted from official air quality monitoring stations aggregated to province level (Regional Environmental Protection Agencies) | Multivariate binomial regression | Daily PM<sub>10</sub> limit exceedances (with a 17 day lag) were significantly associated with COVID-19 cases (coeff=0.25, p<0.001) | N/A | Population density, number of people traveling daily, proportion of people traveling | No |

Table 2 summarizes time-series studies that focus on the short-term effects of air pollution on COVID-19 health outcomes (n=9 studies) (Bashir et al. 2020; Han et al. 2020; Jiang et al. 2020; Li et al. 2020; Wang B et al. 2020; Xu H et al. 2020; Yao et al. 2020a, b; Zhu et al. 2020). These studies compared day-to-day variations in air pollution with day-to-day variations in COVID-19 health outcomes. Table 3summarizes cross-sectional studies that focus on the short-term effects of air pollution on COVID-19 health outcomes (n=8 studies) (Baron 2020; Frontera et al. 2020; Fronza et al. 2020; Setti et al. 2020; Yao et al. 2020a; Yao et al. 2020c; Zhang T et al. 2020; Zhou et al. 2020). These studies compared air pollution levels during the outbreak or shortly before the outbreak with COVID-19 health outcomes. The underlying hypothesis of both time-series and cross-sectional studies is that short-term air pollution exposure increases the transmission of the virus or the severity of COVID-19.

Note that one of the 28 studies included both short-term time-series and short-term cross-sectional study designs (Yao et al. 2020a); this study is included in both Tables 2 and 3.

### Source of the studies

Of the 12 long-term studies, 11 were published in MedRxiv and 1 was published in a peer-reviewed journal. Of the 16 short-term studies, 10 were published in MedRxiv and 6 were published in a peer-reviewed journal.

### Location of the studies

The 28 studies examined the effects of air pollution on COVID-19 health outcomes across 9 different countries (China, Italy, USA, France, Iran, Spain, Germany, United Kingdom, and the Netherlands). Many of the studies focused on cities and provinces in China, regions and provinces in Italy, and counties in the United States, all regions where the SARS-CoV-2 outbreak occurred early and was prominent. Among the short-term studies, 12 were conducted in China, 3 in Italy, 1 in the United States, 1 in France, 1 in Germany, and 1 in Spain. Among the long-term studies, 6 were conducted in Italy, 5 in the United States, 3 in China, 2 in Spain, 2 in France, 2 in Germany, 2 in the United Kingdom, 1 in Iran, and 1 in the Netherlands.

### COVID-19 health outcomes and exposures

5 different COVID-19 health outcomes were included across the 12 long-term studies and 7 different COVID-19 health outcomes were included across the 16 short-term studies. Among the 12 long-term studies, 8 used number of cases, 7 used number of deaths, 1 used case-hospitalizations, 1 used case-fatality rate, and 1 used percent of severe infection. Among the 16 short-term studies, 12 used number of cases, 3 used number of deaths, 2 used case-fatality rate, 2 used basic reproduction number, 1 used ICU admissions, 1 used hospitalized cases, and 1 used a binary variable for epidemic escalation.

The 28 studies also used a variety of air pollution exposures. 11 of the 12 long-term studies collected air pollution data over extended periods of time ranging from 1 to 16 years. These studies used data after 1999 and prior to 2020. One study that self-classified as long-term collected air pollution data from January and February 2020 (Ogen 2020). While this study included air pollution data prior to the outbreak, it does not fully represent long-term exposure, as the data were collected over a period of only 2 months. Ten of the long-term studies obtained air pollution concentrations or air quality index values, and 3 studies used the number of daily limit exceedances.

The short-term time series studies used daily air pollution data collected over relatively shorter periods of time, ranging from 17 days to 80 days. These studies used daily air pollution data during the outbreak with a short lag time to account for incubation period and/or time from exposure to death. However, Bashir et al. (2020) did not discuss the use of a lag period in their analyses, which should be taken into consideration when interpreting the results from that study. Eight of the short-term time-series studies used air pollutant concentrations or air quality index values, and one study used an unspecified air quality metric provided by the National Weather Service (Bashir et al. 2020).

Short-term cross-sectional studies used aggregated air pollution data collected before or during the outbreak. These studies collected air pollution data over periods of time ranging from 1 to 3 months. One of the short-term cross-sectional studies collected air pollution concentration data based on the number of daily limit exceedances over a period of time (Setti et al. 2020). The other 7 studies used air pollutant concentrations or air quality index values.

Overall, 21 studies examined PM_2.5_ (9 long-term, 12 short-term); 16 studied PM_10_ (7 long-term, 9 short-term); 14 studied NO_2_ (7 long-term, 7 short-term); 12 studied O_3_ (8 long-term, 4 short-term); 7 studied CO (3 long-term, 4 short-term); and 7 studied SO_2_ (4 long-term, 3 short-term). Two short-term studies examined Air Quality Index (AQI), 1 long-term study examined NO, 1 long-term study examined aerosols, 1 long-term study examined HCHO, 1 short-term study looked at NH_3_, and 1 short-term study included an unspecified air quality metric provided by the National Weather Service.

### Covariates and Confounders

5 of the 12 long-term studies (Table 1) did not consider covariates and confounders in their analyses. 3 of the 9 short-term time-series studies (Table 2) and 3 of the 8 short-term cross-sectional studies (Table 3) did not consider covariates and confounders in their analyses.

### Statistical models

There were 8 different statistical models used across the 12 long-term studies and 9 different statistical models used across the 16 short-term studies. Among the 12 long-term studies, 3 used linear regression, 2 used negative binomial regression, 2 used negative binomial mixed models, 2 used rank correlation, 1 used descriptive analyses, 1 used non-parametric penalized linear regression, 1 used Poisson regression, and 1 used Partitioning Around the Medoids Clustering Algorithm. Among the 16 short-term studies, 8 used linear regression, 2 used Poisson regression, 2 used rank correlation, 1 used binomial regression, 1 used unspecified analyses, 1 used artificial neural networks (ANN), 1 used negative binomial regression, 1 used a generalized additive model with a Gaussian distribution, and 1 used a generalized additive model with a quasi-Poisson distribution.

### Statistically significant results

Of the 28 studies, 27 showed a statistically significant positive association between air pollution and adverse COVID-19 health outcomes (Figure 2). The only long-term study that did not show a statistically significant association conducted a descriptive analysis of NO_2_ and COVID-19 health deaths (Ogen 2020). All of the 16 short-term studies found statistically significant positive associations between air pollution and adverse COVID-19 outcomes. While 27 of the 28 papers found a statistically significant positive association between an air pollutant and adverse COVID-19 health outcomes, not every air pollutant studied was found to be statistically significant or positively associated with every adverse COVID-19 health outcome studied in every location studied for every lag period analyzed.

**Figure 2.**
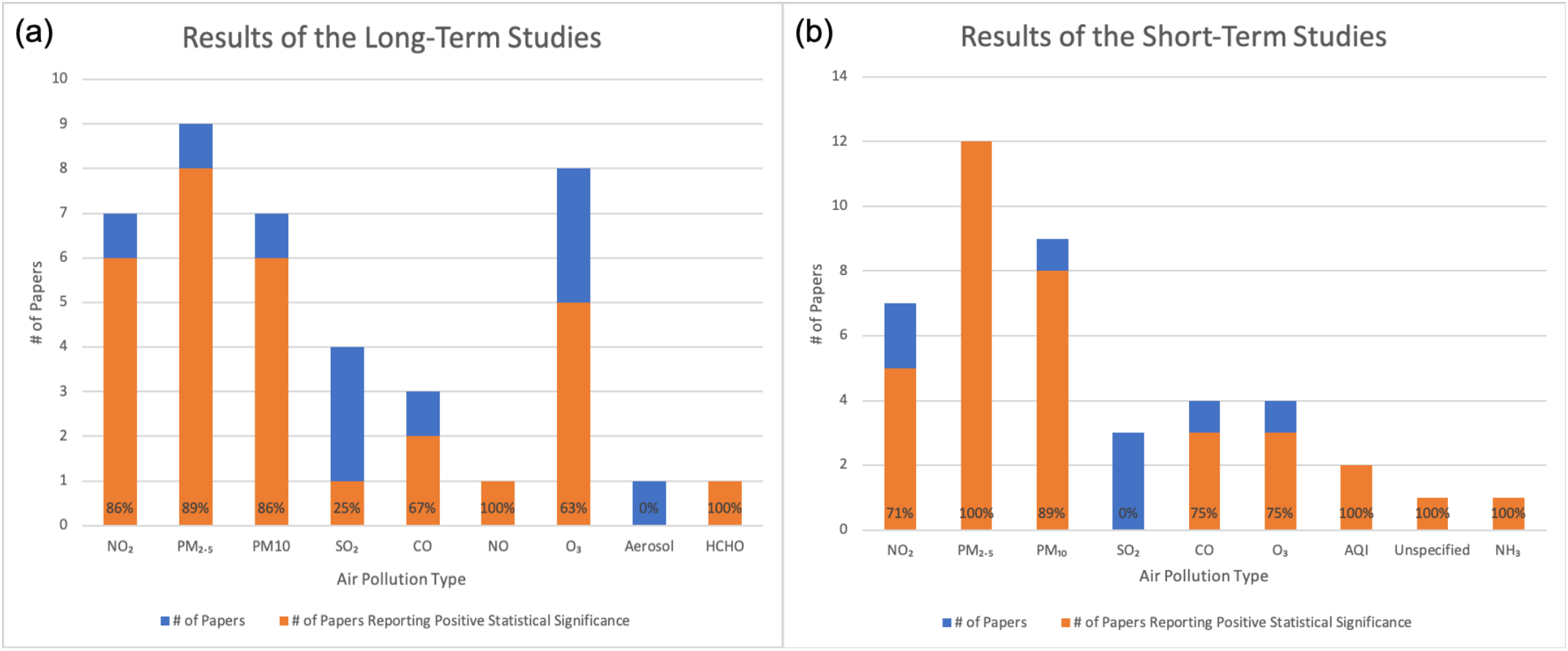
Results stratified by the type of air pollutant. Orange represents the number of studies that reported a statistically significant positive association between the air pollutant and COVID-19 outcomes. Blue and Orange together represent the total number of studies. (a) Long-term studies. (b) Short-term studies.

Among the long-term studies, those that found a statistically significant positive association between a specific air pollutant and adverse COVID-19 health outcomes are as follows: 8 of 9 studies of PM_2.5_; 5 of 8 studies of O_3_; 6 of 7 studies of NO_2_; 6 of 7 studies of PM_10_; 1 of 4 studies of SO_2_; and 2 of 3 studies of CO. The only long-term studies that included HCHO and NO found a statistically significant positive association. The only long-term study of aerosols did not find a statistically significant positive association.

Among the short-term studies, those that found a statistically significant positive association between a specific air pollutant and adverse COVID-19 health outcomes are as follows: all 12 studies of PM_2.5_; 8 of 9 studies of PM_10_; 5 of 7 studies of NO_2_; 3 of 4 studies of O_3_; and 3 of 4 studies of CO. Of the 3 short-term studies of SO_2_, none found a statistically significant positive association with adverse COVID-19 health outcomes, whereas both of the 2 short-term papers studying AQI found a statistically significant positive association. The only papers studying NH_3_ and an unspecified air quality metric provided by the National Weather Service also found statistically significant positive associations with adverse COVID-19 health outcomes.

## Discussion

We searched the literature from January 1, 2020 to June 9, 2020 to identify studies that examined the relationship between air pollution and adverse COVID-19 health outcomes. The query yielded papers published in 2020, and 28 papers satisfied our exclusion/inclusion criteria. These papers used different statistical models, different datasets, and different confounders.

Nonetheless, 27 of the 28 included studies reported some form of statistically significant association between exposure to air pollution and adverse COVID-19 health outcomes.

While researchers want to disseminate their results rapidly to accelerate knowledge about this pandemic as it unfolds, this practice presented us with many challenges when trying to understand the results of the selected studies and their implications. Yet, even in pre-pandemic times, research on the health effects of air pollution is challenging. It is well known that assessing causality in air pollution epidemiology is complicated by the fact that most studies are observational with a large number of confounding factors (Dominici and Zigler 2017). There is also the consistent threat of unmeasured confounding bias. Exposure and outcome misclassification present further challenges.

When the goal is to determine a clear association between air pollution and its effects on health outcomes, such as adverse COVID-19 health outcomes, some well-known challenges become even more serious, and a number of additional complications emerge. These additional challenges are mostly related to the reality that the data on SARS-CoV-2 and COVID-19 are constantly evolving, come from a variety of sources, and are primarily available only at the aggregate level (e.g., county, regional, or country level). Indeed, the quality of the data and the lack of available well-validated electronic health record data at the national level (particularly in the United States) renders the general challenges in evaluating causality enumerated above even more difficult to address. We summarize below the most important methodological challenges, which in turn highlight critically important areas of research that will need to be pursued.

## Data quality

The validity of the health outcome data are questionable, as there is no uniform case definition of a COVID-19 death and there are also diagnostic errors in COVID-19 cases(Gandhi and Singh 2020). This could potentially contribute to a high degree of over- or under-reporting. Many infected people may have died without ever having been tested for SARS-CoV-2, while in other cases, SARS-CoV-2 might have been secondary to the cause of an individual’s death. Moreover, in the early phases of the SARS-CoV-2 pandemic, and even in the current phase of the pandemic, testing was not universally available due to shortages in test kits and the relatively late adoption of mass-scale testing. Additionally, only a subset of the population has been tested, and we cannot account for those who are asymptomatic but carry the virus.

## Ecological fallacy

27 of the 28 studies included were ecological studies as opposed to individual-level studies. The only study conducted at the individual level was conducted by a group in England, where links between air pollution and adverse COVID-19 health outcomes were studied by looking at patient health records (Travaglio et al. 2020). Ecological fallacy is a formal fallacy in the interpretation of statistical data that occurs when inferences about the nature of individuals are deduced from inferences about the group to which those individuals belong (see for example (Jackson et al. 2008)). While ecological studies are often very useful at generating preliminary evidence in data-scarce settings, increasing the scientific rigor of research in this area requires access to nationally representative, individual-level data on adverse COVID-19 health outcomes, including information about patients’ residential address, demographics, and individual-level confounders. This is an enormous challenge that will require many privacy, legal, and ethical trade-offs (Sittig and Singh 2020).

## Other determinants of COVID-19

Ecological data typically do not allow for adjustment by individual risk factors such as age, gender, ethnicity, or occupation. Age is one of the strongest predictors of survival for most conditions, including COVID-19. Gender-based differences in time-activity patterns contribute to different levels of air pollution exposure between men and women, and women have been shown to be more susceptible to several environmental exposures (Clougherty 2010). Occupation is also an important factor in this pandemic, as those who provide medical care and other essential workers, such as those working in meat packing plants, are at increased risk of developing SARS-CoV-2 infection (Mutambudzi et al. 2020).

## Accounting for difference and socioeconomic status

It is also important to recognize that disadvantaged people (e.g., those without health insurance, undernourished, and with poorly managed underlying health conditions, such as cardiovascular conditions and/or diabetes) have a greater susceptibility for both contracting SARS-CoV-2 and dying from COVID-19. Myriad social and economic factors contribute to high rates of infection and put individuals at higher risk from the sequelae of COVID-19, and disparities in COVID-19‒related outcomes may be due to social deprivation rooted in long-standing racial and socio-economic inequities. This issue has been raised by a number of authors (see for example (Chowkwanyun and Reed 2020; Yancy 2020)).

## Exposure error

In most of the studies reviewed, both long-term and short-term, the same level of air pollution exposure was assigned to everyone living in large geographical areas. Therefore, spatial differences in exposure were not captured. Several statistical approaches have been developed to propagate the different sources of uncertainty associated with exposure error into the statistical model to estimate health effects (see for example (Dominici et al. 2000; Gryparis et al. 2009; Szpiro et al. 2011)). These approaches have not yet been implemented in studies of air pollution exposure and adverse COVID-19 health outcomes.

## Physical distancing

The implementation of public health policies, which can vary widely by jurisdiction, has been shown to be successful in reducing SARS-CoV-2 transmission and flattening the epidemic curve. For example, in Georgia, areas that did not adopt physical distancing practices experienced higher incidence and mortality from COVID-19 when compared to other areas in the state that did. Cities in California that tend to have higher levels of fine particulate matter adopted stay-at-home policies earlier than other regions. Because these policies differ by regional air pollution levels, including rural and urban areas within the same county, they can distort the observed associations between air pollution and adverse COVID-19 health outcomes.

## Timing on the epidemic curve

There will be temporal differences in the number of incident COVID-19 cases, and by extension in COVID-19‒related deaths, by region. US counties were at very different stages on the epidemic curves, especially in early April. Larger cities are more populous and tend to have increased travel to and from international locations, providing increased opportunity for the spread of COVID-19 early in the pandemic. These larger cities also tend to have higher concentrations of air pollution. The practical implications are that there will be a greater number of incident cases and deaths in those cities that are further along on the epidemic curve, which further confounds analysis.

## Clustering of cases and deaths

Unlike studies of long-term exposure to air pollution and chronic diseases where deaths can reasonably be assumed to be independent, COVID-19 cases and deaths tend to occur in clusters. This has been widely reported, such as the now famous choir practice where a large portion of attendees became ill or the tragic events in congregant settings such as retirement homes and long-term care facilities. Although some of the studies’ authors included random effects to account for clustering, without individual-level data, it is simply not possible to account for this clustering.

## Reproducibility

To discover crucial linkages between air pollution and adverse COVID-19 outcomes in a more definitive, causal manner, both the data used for the analyses as well as the code should be made publicly available. Transparency and shared resources will assist in the global push towards uncovering the solutions to the pandemic and instituting public policies that will protect the health of people worldwide. Only two of the 28 selected papers included publicly-available code. Many of the selected papers were in preprint format, and access to code will be critical to validate results and build upon the conclusions. It is difficult to reproduce results and continue work on these areas without code availability.

## Conclusion

Assessing the short- and long-term effects of air pollution on adverse COVID-19 health outcomes is a rapidly evolving area of research. Most of these studies are still in the preprint stage and many more studies will likely be published in the next few weeks and months. Therefore, our aim was to identify this topic as a critically important area of research, identify the numerous methodological challenges, and inform future research. Despite the preliminary nature of the evidence summarized in this scoping review, our findings underscore the need to hold governments accountable for the installation of environmental protections that will permanently maintain safe levels of air pollution to protect human health, rather than removing those environmental protections at the behest of the industries that pollute our environment. In the era of climate change, and now global pandemics, studies like these are among many calling for action to protect the earth systems that are so deeply intertwined with human systems, particularly systems of human health.

A primary goal of this scoping review was to begin paving the way toward overcoming the many methodological challenges that are inherent in studies of environmental health epidemiology. Some of these challenges are common to all epidemiological studies of air pollution and health outcomes (e.g., exposure error, measured and unmeasured bias), while others are exacerbated in the study of adverse COVID-19 health outcomes (e.g., outcome misclassification, evolving number of new cases and deaths) and global pandemics (e.g., accounting for different testing practices, different stages of the pandemic).

With respect to these new challenges, it is just as important to highlight new opportunities. For example, the extreme measures implemented during the lockdown are providing new research opportunities to investigate important questions regarding achievable reductions in air pollution exposure and health effects. Indeed, in order to mitigate the effects of the pandemic and to protect lives, countries across the globe instituted temporary, and in many cases, continued closures of all but essential businesses and services, instituting lockdowns to encourage people to stay at home and prevent the spread of disease. This has resulted in significantly lower levels of vehicular, train, aircraft, and industry related emissions (Le Quéré et al. 2020; Shilling and Waetjen 2020; Zhang R et al. 2020). These lockdown measures provide a unique opportunity to exploit the features of a quasi-experimental design to assess the extent to which different pollutants have declined and estimate the potential “beneficial” effects of these declines on health outcomes.

## Data Availability

N/A

## Acknowledgements

We gratefully acknowledge support from the 2020 Star-Friedman Challenge for Promising Scientific Research which supported Dr. Dominici and Dr. Braun.

## Supplemental Materials

**Supplemental Table 1.**
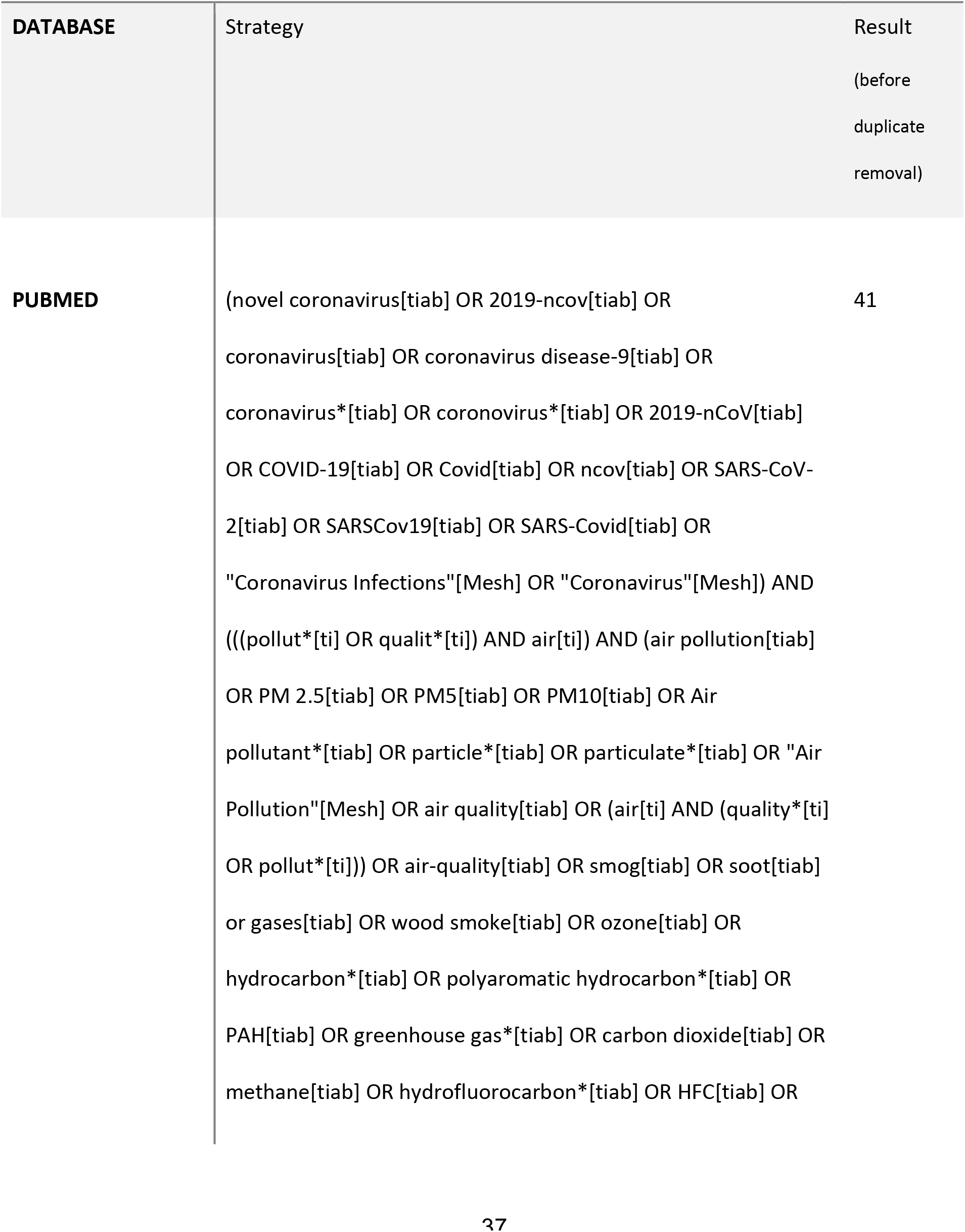
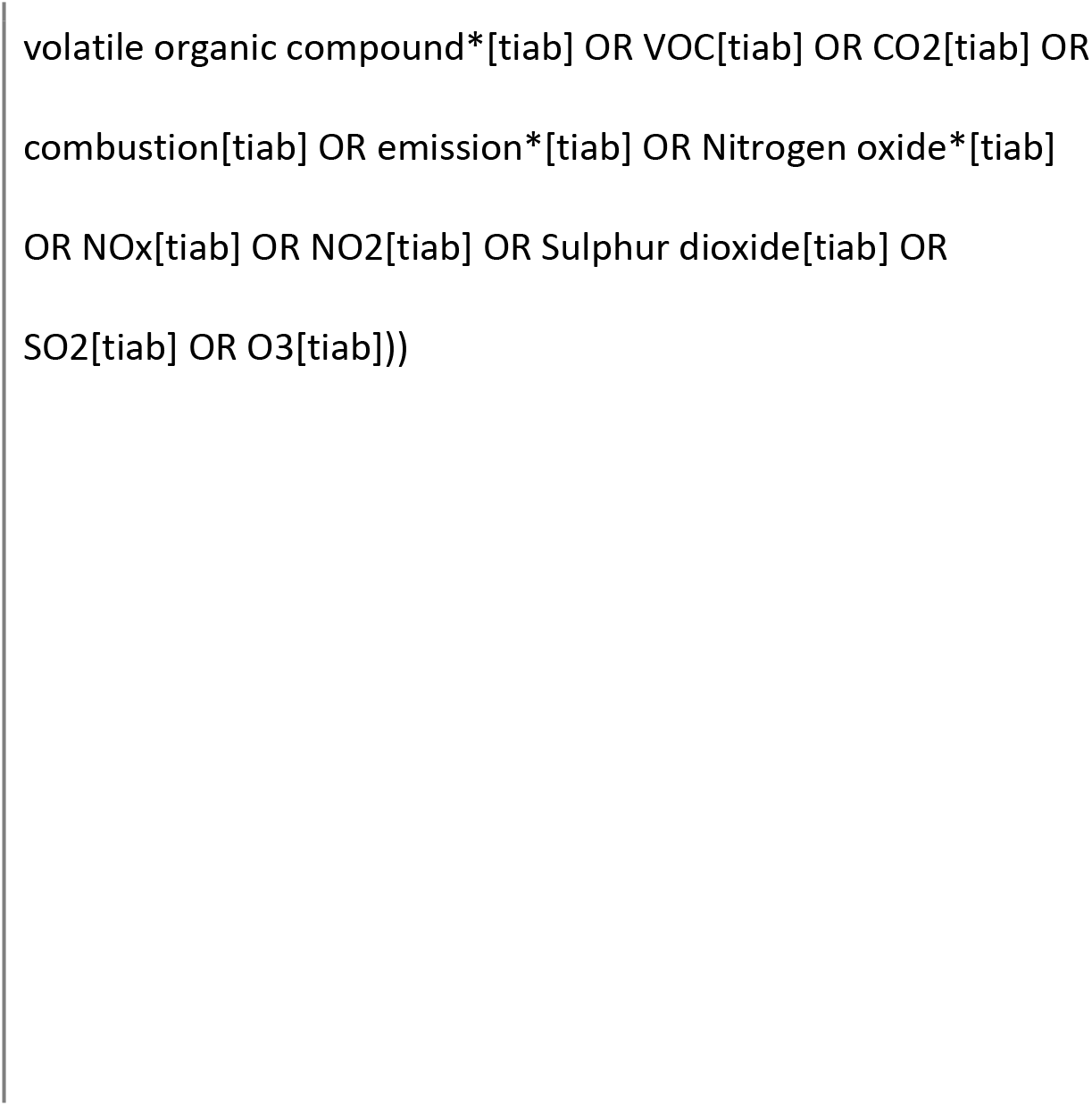
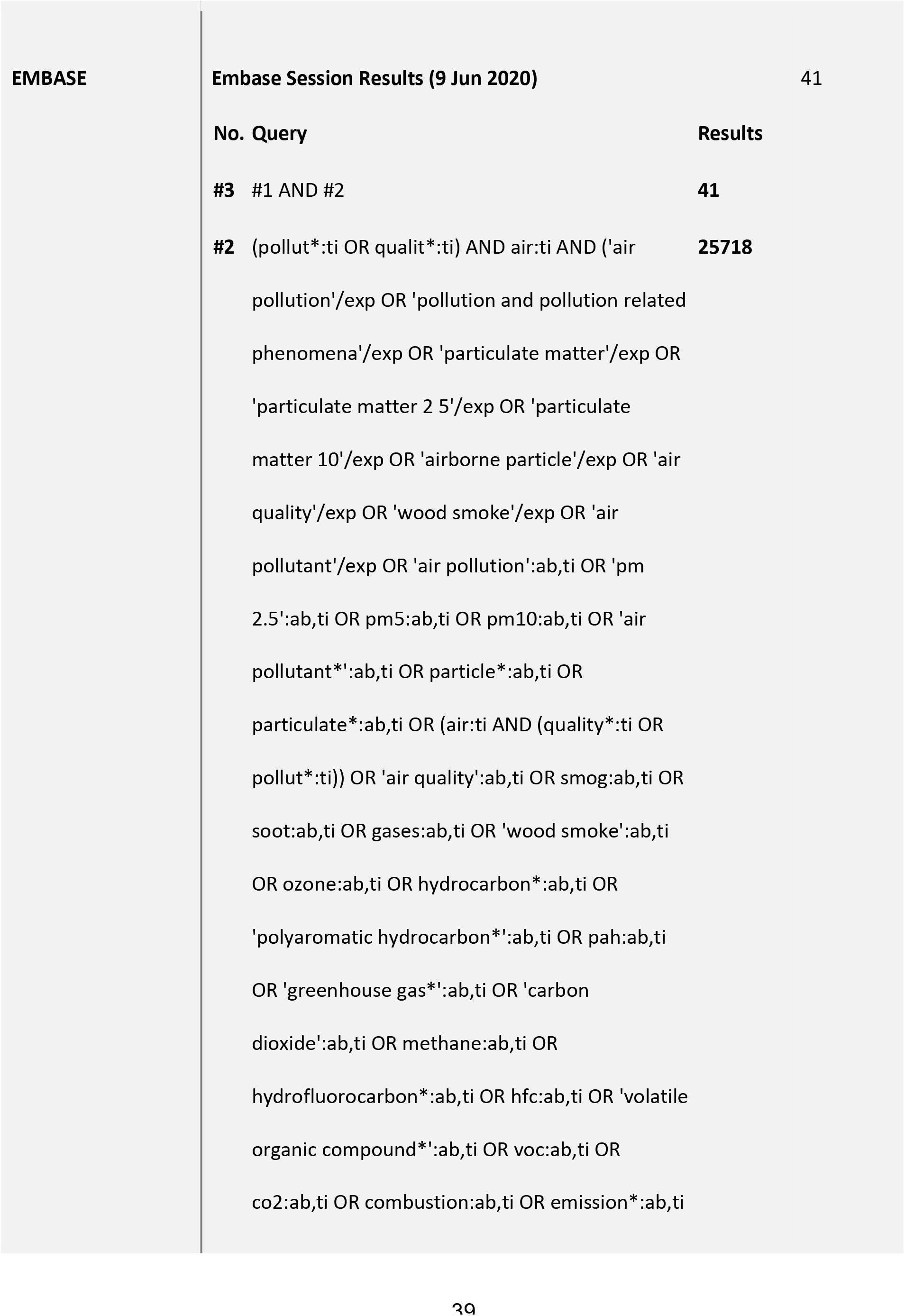
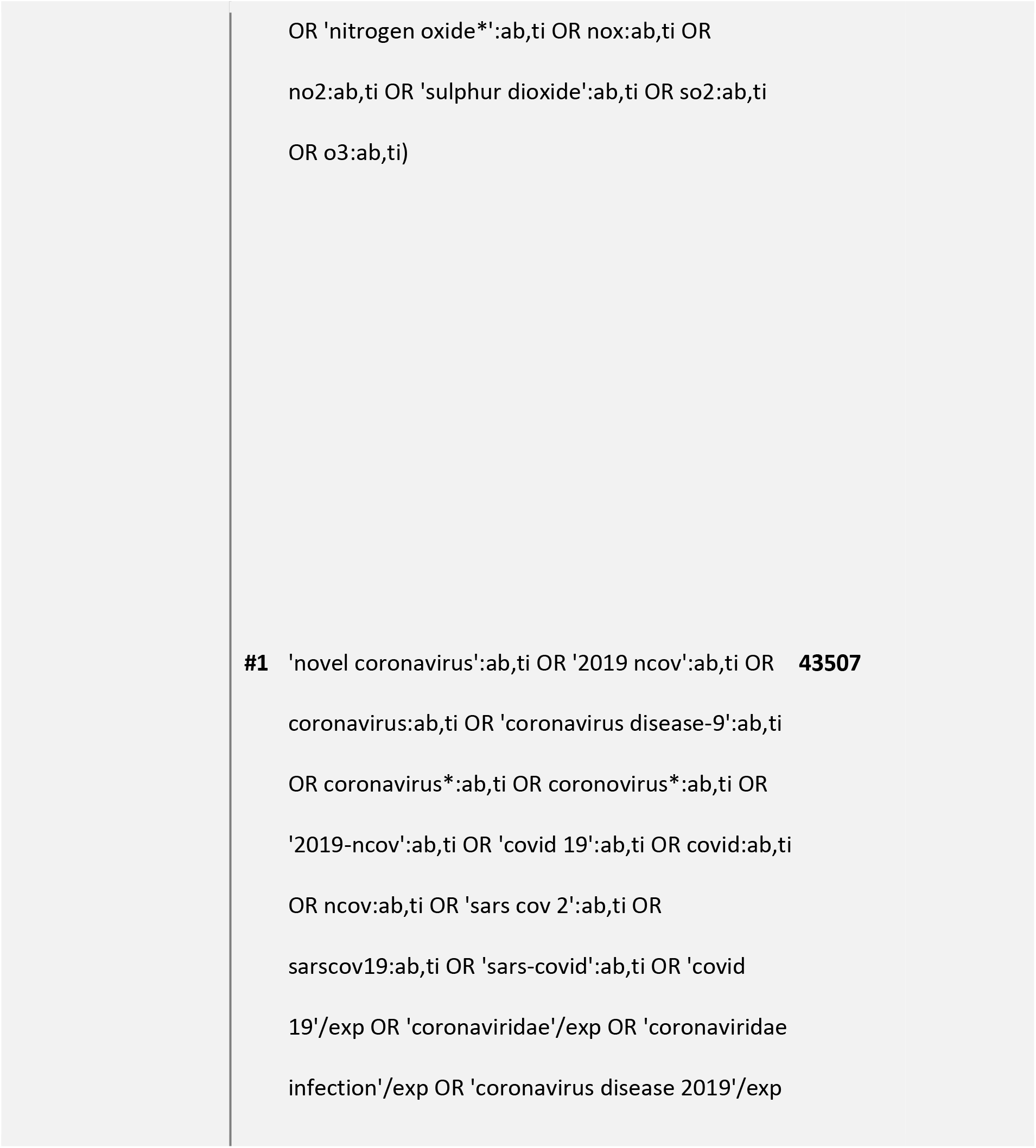
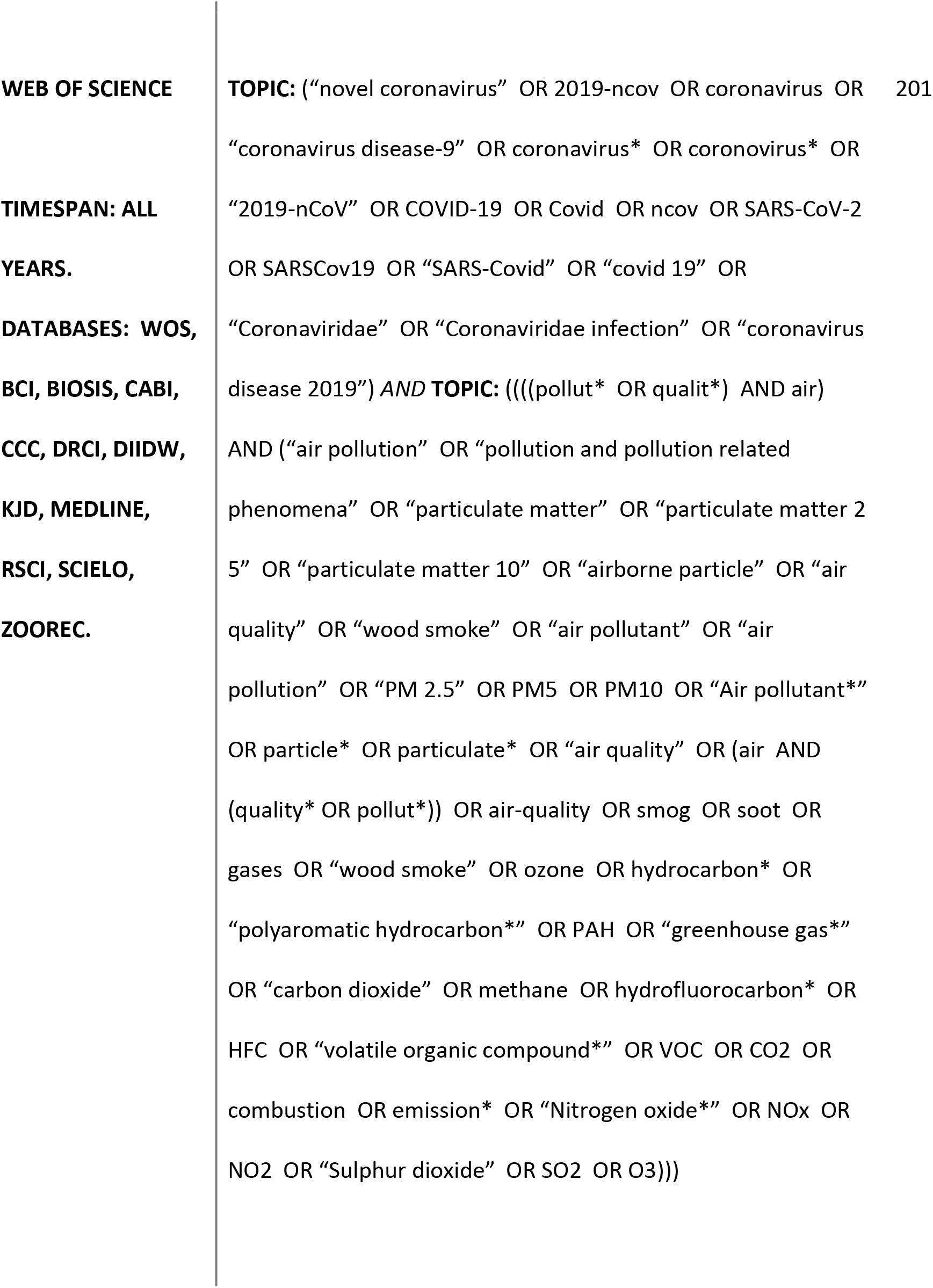
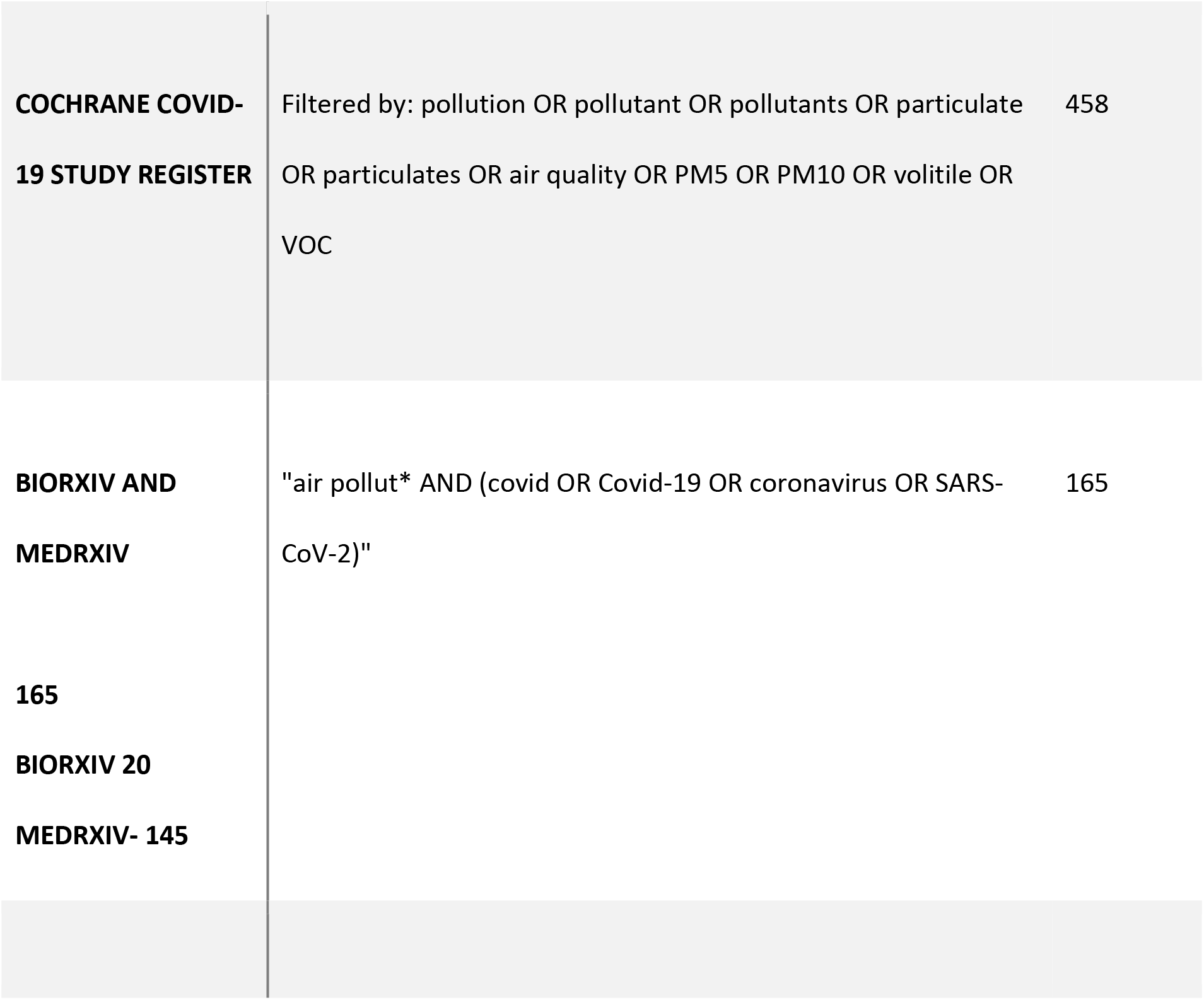
Supplemental Table 1. Search Queries for All Relevant Databases (as of June 9, 2020)

